# Distal Ventricular Pacing for Drug-Refractory Mid-Cavity Obstructive Hypertrophic Cardiomyopathy: A Randomized, Placebo-Controlled Trial of Personalized Pacing

**DOI:** 10.1101/2023.12.18.23300178

**Authors:** James W Malcolmson, Rebecca K Hughes, Tim Husselbury, Kamran Khan, Annastazia E Learoyd, Martin Lees, Eleanor C Wicks, Jamie Smith, Alexander Simms, James Moon, Luis Lopes, Constantinos O’Mahony, Neha Sekhri, Perry Elliott, Steffen E. Petersen, Mehul Dhinoja, Saidi A Mohiddin

**Author notes:** **Corresponding author:** Professor Saidi. A. Mohiddin Department of Inherited Cardiovascular Disease Unit SBH St Bartholomew’s Hospital West Smithfield London EC1A 7BE.

## Abstract

**Background:** Patients with refractory symptomatic left ventricular (LV) mid-cavity obstructive (LVMCO) hypertrophic cardiomyopathy (HCM) have few therapeutic options. Right ventricular (RV) pacing is associated with modest hemodynamic and symptomatic improvement, and LV pacing pilot data suggest therapeutic potential. We hypothesized site-specific-pacing would reduce LVMCO gradients and improve symptoms.

**Methods:** Patients with symptomatic-drug-refractory LVMCO were recruited for a randomized blinded trial of personalized prescription of pacing (PPoP). Multiple LV and apical RV pacing sites were assessed during invasive hemodynamic study of multisite pacing. Patient-specific pacing-site and atrioventricular (AV) delays, defining PPoP, were selected on basis of LVMCO gradient reduction and acceptable pacing parameters. Patients were randomized to 6-months of active PPoP or back-up pacing in a cross-over design. The primary outcome examined invasive gradient change with best-site pacing. Secondary outcomes assessed quality of life and exercise following randomization to PPoP.

**Results:** A total of 17 patients were recruited; 16 of whom met primary endpoints. Baseline NYHA was 3±0.6 despite optimal medical therapy. Hemodynamic effects were assessed during pacing at the RV apex and at a mean of 8 LV sites. The gradients in all 16 patients fell with pacing, with maximum gradient reduction achieved via LV pacing in 14 (88%) patients and RV apex in 2. The mean baseline gradient of 80±29 mmHg, fell to 31±21 mmHg with best-site pacing, a 60% reduction (p<0.0001). One cardiac vein perforation occurred in one case, and 15 subjects entered cross-over; 2 withdrawals occurred during cross-over. Of the 13 completing cross-over, 9 (69%) chose active pacing in PPoP configuration as preferred setting. PPoP was associated with improved 6-minute walking test performance (328.5±99.9 vs 285.8±105.5 meters, p=0.018); other outcome measures also indicated benefit with PPoP.

**Conclusions:** In a randomized placebo-controlled trial, LV pacing reduces obstruction and improves exercise performance in severely symptomatic LVMCO patients.

**Registration:** NCT03450252.

**Clinical Perspective:** *What is Known?:*

- Patients with refractory, symptomatic LVMCO present a significant challenge for clinical management, with very few treatment options.
- Data on the use of right ventricular (RV) pacing in patients with refractory, symptomatic LVMCO indicate suboptimal therapeutic responses whilst pilot data indicate a potential therapeutic role for LV pacing.

*What the Study Adds?:*

- Personalized prescription of pacing (PPoP) therapy guided by invasive hemodynamics significantly reduced LVMCO gradients and improved exercise performance in the first randomized, placebo-controlled trial in symptomatic LVMCO.
- This study provides the basis for a multicenter trial of PPoP for LVMCO and for the use of site-specific pacing in managing other forms of HCM.

**Graphical Abstract:** 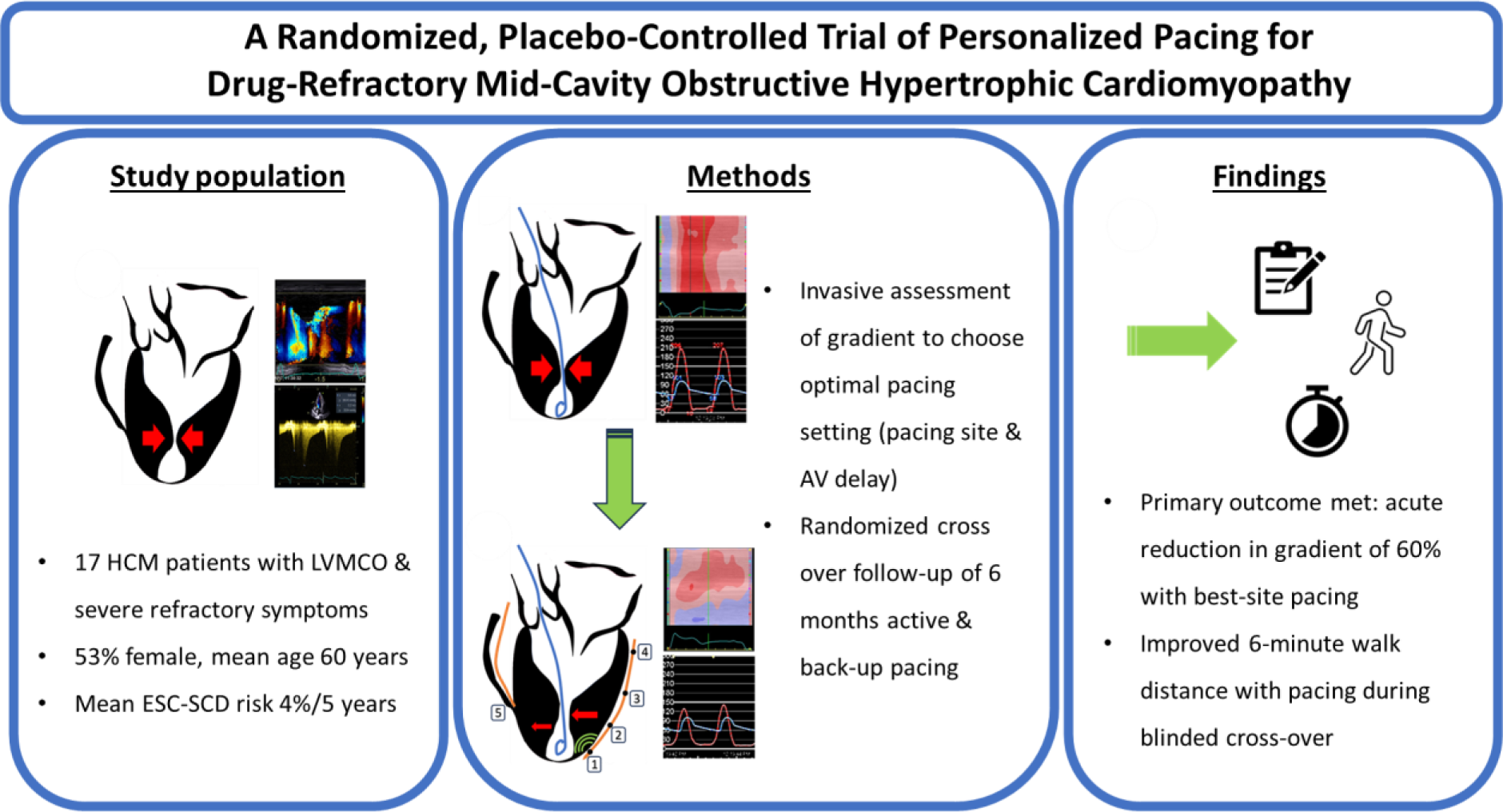

## 1. Introduction

Hypertrophic cardiomyopathy (HCM) is the most common inherited heart disease, affecting around 1 in 500 people.^1^ HCM is characterized by abnormal myocardial thickening and hypercontractility, with obstruction to blood flow within the left ventricle (LV) frequently seen at the level of the outflow tract, associated with morbidity and adverse prognosis.^2, 3^ LV mid-cavity obstruction (LVMCO) is a less commonly recognized phenotypic form of HCM in which obstruction during LV ejection occurs due to partial or complete obliteration and division of the LV cavity into two distinct areas (basal and apical). Here, hyperdynamic muscular contraction forms a constricting muscular neck at the point of cavity division.^4^ ^5^ High intracavity pressure gradients form across the point of obstruction, often associated with the development of discrete LV apical aneurysms (LVAA),^6^ a risk factor for adverse events.^7^

Patients with LVMCO are often symptomatic,^8^ and first-line pharmacologic therapies (including betablockers, calcium channel antagonists, and disopyramide) aim to reduce LV inotropy and/or increase filling time. Those with symptoms refractory to medical therapy have severely limited therapeutic options. Studies have indicated that pacing may reduce obstructive gradients and improve symptoms in LVMCO.^9, 10^ Notably, as we and others have shown,^4, 11, 12^ Doppler derived assessments often severely underestimate LVMCO gradient magnitude and invasive methods are needed for accurate measurement.

Our group has pioneered the use of invasive hemodynamic assessments made during a multi-site pacing study to determine optimal pacing configurations. Initial data obtained in severely symptomatic LVMCO patients with conventional indications for device therapy demonstrated that this personalized prescription of pacing (PPoP) approach reduced LVMCO gradients, and was associated with symptomatic improvement in an unrandomized cohort of 16 patients.^13^ These observational data informed the design of this randomized, placebo controlled cross-over trial of PPoP. Here, we hypothesized that PPoP as guided by invasive hemodynamic measurements of arterial and LV pressures to accurately define obstructive gradients^4^ would reliably reduce LVMCO gradient severity (primary aim) and improve functional status (secondary aims).

## 2. Methods

### Study design and participants

This was a single-center, prospective study of distal ventricular pacing for gradient reduction and symptomatic relief in HCM patients with isolated LVMCO. Ethical approval was granted by the National Research Ethics Service, City Road and Hampstead, London (Reference: 17/LO/1725). The trial was performed in agreement with the Declaration of Helsinki and registered with the following public registries: http://clinicaltrials.gov (NCT03450252) and International Standard Randomized Controlled Trials Number (ISRCTN): ISRCTN82621856. The study was funded by the National Institute for Health and Care Research (NIHR) in the United Kingdom.

Only LVMCO patients with severe drug-refractory symptoms were included. Eligible HCM patients were ≥18 years with LVMCO gradient ≥30 mmHg demonstrated initially by echocardiography, and confirmed by cardiac catheterization at rest or with isoprenaline provocation, referred for pacemaker (PPM) +/- implantable cardioverter defibrillator (ICD) implantation for either primary prevention of sudden cardiac death or other indications such as heart block or obstructive physiology; patients were taking maximum tolerated doses of beta blockers or verapamil +/- disopyramide.

Exclusion criteria included multi-level obstruction (i.e. across the mid-cavity and outflow tract if the latter was determined to be the dominant lesion); moderate or severe primary valvular disease; untreated symptomatic coronary disease; atrial fibrillation at the time of device implantation; pregnancy; eGFR <20mL/min; and patients unable to provide informed consent. Inclusion and exclusion criteria are fully listed in Table 1.

**Table 1:**
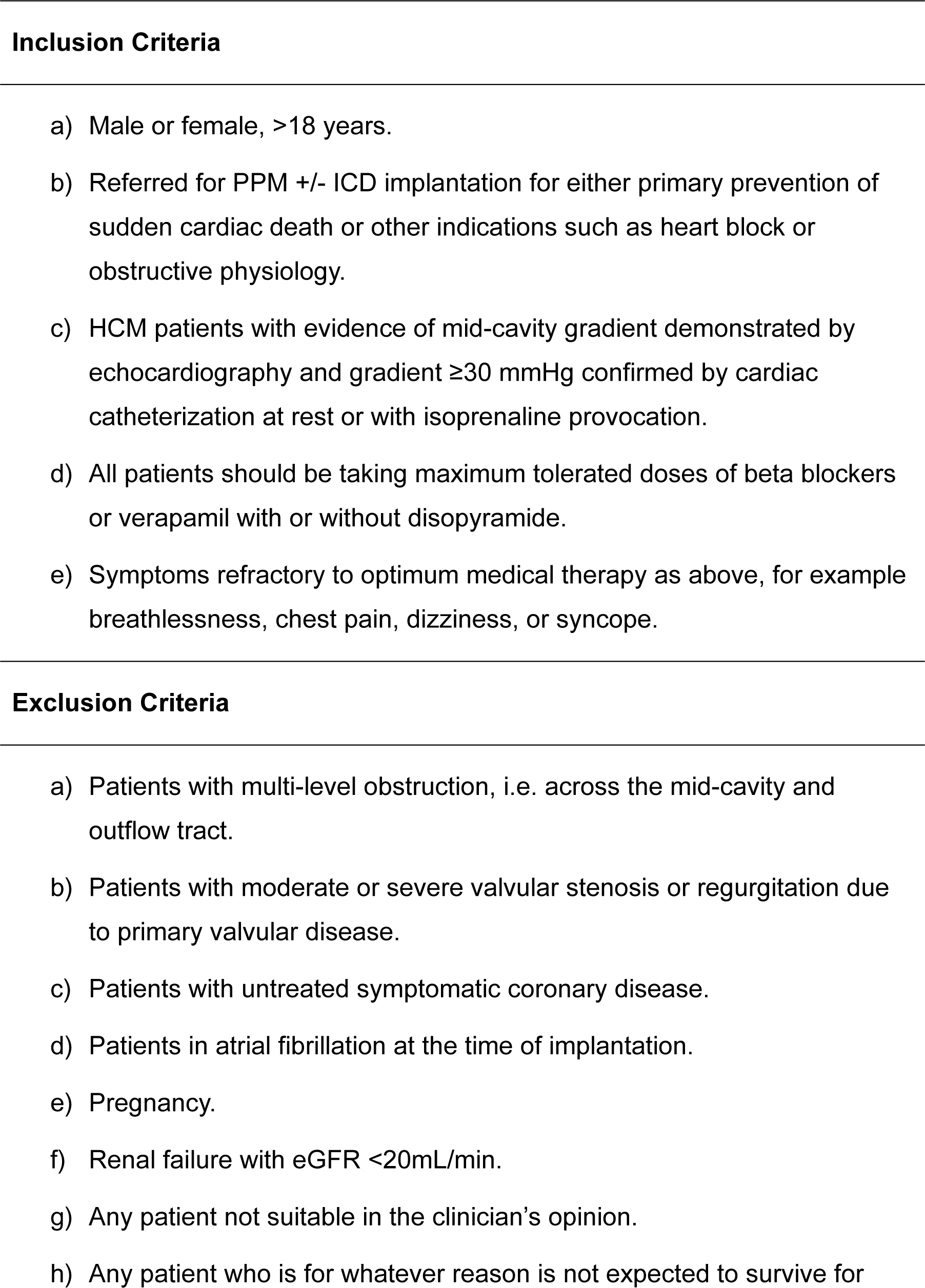
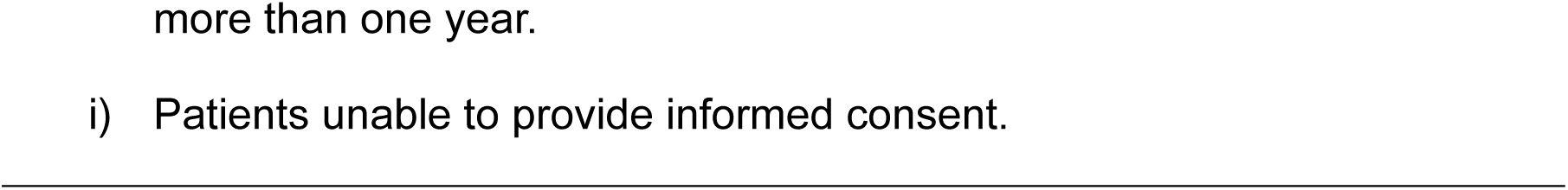
Inclusion and exclusion criteria.

### Study design overview

Baseline evaluation of symptom and performance (secondary outcome measures) included NYHA class, Short Form-36 (SF-36) questionnaire scores, Kansas City Cardiomyopathy Questionnaire (KCCQ) scores, 6-minute walk test (6MWT) distance, cardiopulmonary exercise testing (CPET) with stress echocardiography, and serum concentration of N-terminal pro B-type natriuretic peptide (NT-proBNP).

Subjects then underwent an invasive hemodynamic assessment of multi-site pacing study to guide LV and RV lead positioning and device implantation. Primary outcome data were collected during the hemodynamic pacing study and comprised of the acute invasively determined changes in LVMCO gradient at the optimal pacing site when compared to sinus rhythm. Quadripolar LV lead position was determined according to a pragmatic synthesis of pacing-site-specific hemodynamic data and appropriate pacing parameters at that site (including thresholds and diaphragmatic/phrenic capture).

On the day following device implant, participants were randomized to either active or back up pacing for the first phase of follow-up (6 months), before crossover to the alternate setting for the second 6-month period. Secondary outcome data were again assessed at the end of each follow-up phase. Participant and Principal Investigator were blinded to pacing status, and the subject’s stated preference of either 1^st^ or 2^nd^ crossover phase was a prespecified secondary outcome measure.

### Hemodynamic pacing study

We have previously described the technique for hemodynamic pacing procedure in detail.^13^ Briefly, all hemodynamic pacing studies and device implants were performed in a single procedure under general anesthesia (GA). Arterial access was achieved via the right femoral artery using the Seldinger technique (operator 1) with central venous access similarly achieved via the left sub-clavian or cephalic veins (operator 2).

Operator 2 advanced the right atrial and RV leads to the heart via the superior vena cava, where they were implanted using standard techniques for active leads (manufacturer dependent). The coronary sinus was intubated using a LV lead delivery guiding catheter and the coronary venous anatomy defined by simultaneous balloon occlusion coronary sinus venography and LV angiography. The coronary vein of interest was intubated with a deflectable quadripolar catheter.

During the hemodynamic pacing protocol, pressures were transduced simultaneously from the LV apex using a specially manufactured end-hole pigtail catheter (Cordis^TM^) and femoral artery via a 7French side arm sheath. Baseline peak-to-peak obstructive gradients were calculated in sinus rhythm; if no resting obstructive gradient was present under GA conditions, steady state isoprenaline infusion was used in provocation: this began at 1 microgram/minute and, if required, was gradually up-titrated to a maximum of 4 micrograms/minute.^14^ ^15^

To select the optimal sensed AV delay, an initial AV delay (60 ms) was lengthened until evidence of QRS fusion was seen on the surface electrocardiogram (ECG) and/or when acute gradient reduction was lost. For each of several pacing configurations (quadripolar LV catheter, RV apex; in unipolar and bi-polar) the best ventricular pacing location (primary outcome) was identified solely according to the greatest acute reduction in obstructive gradient. Each pacing site was systematically tested by turning pacing ‘on’ for a period of 30 seconds to allow stabilization of hemodynamics, before averaging invasively defined LVMCO gradients over 3 consecutive cardiac cycles. When pacing was turned ‘off’ at an individual site, another 30 second period was initiated to allow for stabilization of intracardiac hemodynamics. When selecting the pragmatic pacing site for PPoP, other pacing related factors (including capture threshold, r-wave sensitivity, diaphragmatic capture, change in surface ECG QRS morphology, lead stability) and hemodynamic responses (the maintenance or improvement in systolic arterial pressure) were also considered by the Principal Investigators (operators 1 and 2).

The assessment of obstructive gradients during various pacing configurations was obtained sequentially, beginning with single site pacing from the RV apex, followed by multi-site LV pacing from a quadripolar lead in the middle cardiac or other cardiac vein (venous anatomy dependent). Hemodynamic consequences of multi-site LV pacing were assessed sequentially from distal to proximal poles 1-4. Repositioning of the quadripolar lead in the same or alternative cardiac veins was performed when sub-optimal hemodynamic results were obtained, and/or when pacing parameters or lead stability were unsatisfactory. After final pacing parameters were identified, careful pull-back of the pigtail catheter confirmed site and magnitude of resting and pacing obstruction within the LV.

### Statistical analysis

#### Sample size calculation

Effect size was calculated using our retrospective LVMCO pacing data,^13^ where mean acute reduction in mid-cavity gradient with distal ventricular pacing was 60±26 mmHg. A conventional significant obstructive gradient is 30 mmHg,^16^ and using a conservative approach, we aimed to be able to detect a reduction of 25 mmHg. With the two-sided alpha level set at 0.05 a priori, and a power of 90%, the calculated sample size using a paired sample t-test was 15.

#### Randomization

Patients were randomized to either active or back-up pacing one day after device implantation using a 1:1 ratio. A master randomization list was generated in an appropriate statistical package (STATA, using the ralloc command) to active or back-up pacing with block size varied randomly.

#### Analysis of primary endpoint

All patients completing the initial hemodynamic pacing study were eligible for primary analysis. Comparison of the gradients during ventricular pacing and sinus rhythm was made using a paired t-test. The effect size is presented as the mean difference (optimum pacing minus sinus rhythm gradients) and 95% confidence interval. In addition, the mean values and standard deviations (SD) for gradients during both sinus rhythm and pacing are presented.

#### Analysis of secondary endpoints

The proportions completing each assessment and the proportion withdrawing were used to assess feasibility of the study. Secondary outcomes were compared between baseline and pacing settings using repeated measures analyses (ANOVA or Friedman tests depending on normality of distribution using the D’Agostino & Pearson test) with multiple comparisons tests (Tukey’s or Dunn’s depending on normality) providing a direct comparison between active and back-up pacing settings. Changes between pacing settings during randomized follow-up in each variable were calculated as the value for active pacing minus the value for back-up pacing for each patient. Data are presented as mean±SD or median and interquartile range (IQR) as appropriate.

Data was examined for the presence of carry-over period effects. No carry-over effect was found. There was minor evidence of a period effect influencing the KCCQ results which was not deemed to substantially affect the results presented. All analyses were performed in R^TM^ version 2022.27.1, and figures created in Graphpad Prism^TM^ version 9.5.1.

#### Study Overview and Data Monitoring

Study overview was provided by the Cardiovascular Clinical Trials Unit (CVCTU) at the William Harvey Research Institute, London. Trial safety data was reviewed routinely by an independent Data Safety Monitoring Committee, and oversight provided by independent Trial Steering Committee. All trial data was held in a secure database (REDCAP^TM^), source data verification undertaken by CVCTU monitors, and statistical analyses performed by CVCTU statistician.

## 3. Results

### General characteristics of population

Between February 2018 and March 2022, 17 patients were recruited to the trial. 29 patients were pre-screened for eligibility during work-up for potential invasive therapy for LVMCO and refractory symptoms in a specialist heart muscle / electrophysiology clinic after referral from their primary clinician (Figure 1). Of these 29, 12 patients were excluded after not meeting symptomatic inclusion criteria (n=11), and 1 clinical event prior to trial recruitment taking place (symptomatic ventricular arrhythmia and urgent dual chamber ICD implant).

**Figure 1:**
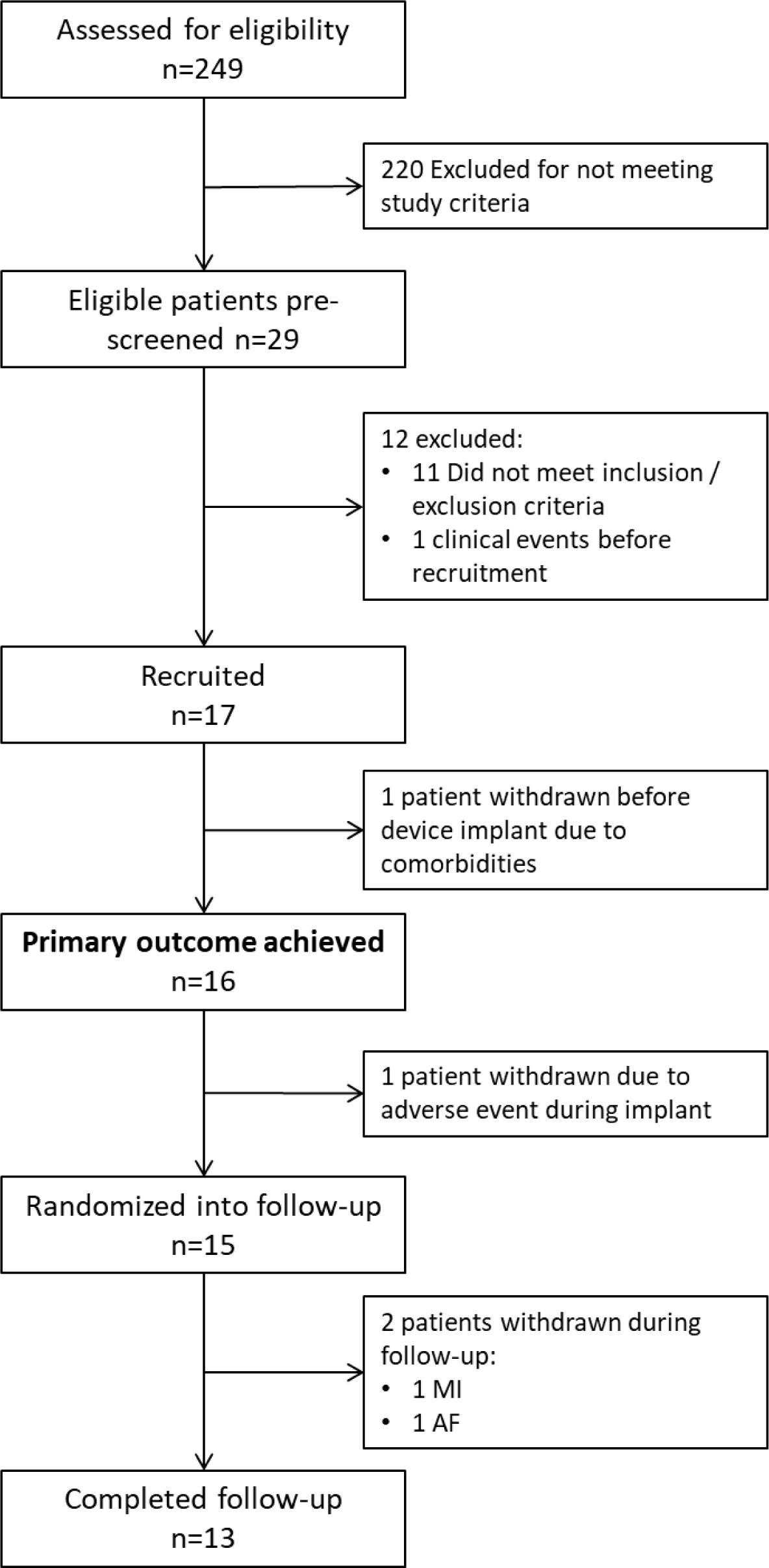
Trial consort diagram. (n, number; MI, myocardial infarction; AF, atrial fibrillation)

### Baseline characteristics

The study population consisted of 17 patients on maximal tolerated medical therapy with LVMCO. Of these, 53% were female, and mean age at recruitment 55.9±10.3 years (Table 2). One patient was withdrawn before the implant procedure due to comorbidities. Of the 16 who underwent hemodynamic pacing study and had devices implanted, 15 (94%) patients received an ICD and one patient received a pacemaker device. At baseline assessment, 17 (100%) patients reported exertional dyspnea, 15 (88%) reported exertional chest pain, 16 (94%) reported palpitations, and 14 (82%) reported presyncope. Seven patients (41%) reported prior unexplained complete loss of consciousness.

**Table 2:** Baseline characteristics.

| **Demographics and symptoms** | **n=17** |
| --- | --- |
| Age at recruitment (years) | 55.9 ± 10.3 |
| Male, n (%) | 9 (53) |
| Family history inherited heart disease, n (%) | 10 (59) |
| HTN, n (%) | 11 (65) |
| Chest pain, n (%) | 15 (88) |
| Dyspnea, n (%) | 17 (100) |
| Palpitations, n (%) | 16 (94) |
| Presyncope, n (%) | 14 (82) |
| **SCD risk profile** | **n=17** |
| Family history of SCD, n (%) | 1 (6) |
| Unexplained syncope, n (%) | 7 (41) |
| Prior NSVT on Holter / ICD (out of 16), n (%) | 09/16 (56) |
| Maximum LV wall thickness ≥30 mm, n (%) | 0 (0) |
| LVAA, n (%) | 14 (82) |
| LVOTO gradient ≥30 mmHg, n (%) | 0 (0) |
| ESC SCD risk score (% 5-year mortality) | 3.7 ± 2.1 |
| ≥ intermediate risk score, n (%) | 6 (35) |
| SCD risk factors (0/1/2/3 risk factors), n (%) | 2 (12) / 9 (53) / 5 (29) / 1 (6) |
| **Medications** | **n=17** |
| β-Blockers, n (%) | 9 (53) |
| Calcium channel blockers, n (%) | 13 (76) |
| Disopyramide, n (%) | 6 (35) |
| Anticoagulation, n (%) | 7 (41) |
| Number on 1/2/3 medical therapies, n (%) | 9 (53) / 7 (41) / 1 (6) |
| **Echocardiography** | n=17 |
| Max LVWT (mm) | 20 ± 4 |
| Resting Doppler LVMCO gradient (mmHg) | 32 ± 21 |
| Post exercise Doppler LVMCO gradient (mmHg) | 50 ± 35 |
| LA diameter (mm) | 40 ± 5 |
| LVAA, n (%) | 14 (82) |
| Paradoxical apical diastolic flow, n (%) | 13 (76) |
| **CMR** | n=12 |
| LVEF (%) | 69 ± 9 |
| Max LVWT (mm) | 20 ± 3 |
| LV mass (g) | 167 ± 37 |
| LVAA, n (%) | 9 (75) |
| Presence of LGE, n (%) | 12 (100) |
| Apical LGE, n (%) | 12 (100) |
| Circumferential perfusion defect (out of 9), n (%) | 8/9 (89) |
| Apical thrombus, n (%) | 0 (0) |

Data are represented as mean ± SD or n (%). HTN, hypertension; SCD, Sudden Cardiac Death Risk; NSVT, non-sustained ventricular tachycardia; ICD, implantable cardioverter defibrillator; LV, left ventricular; LVAA, left ventricular apical aneurysm; LVOTO, left ventricular outflow tract obstruction; ESC, European Society of Cardiology; LVWT, LV wall thickness; LVMCO, left ventricular mid-cavity obstruction; LA, left atrial; CMR, cardiac magnetic resonance; LVEF, left ventricular ejection fraction; LGE, late gadolinium enhancement.

### Primary end point: acute change in invasive gradient

Primary end point assessment was completed in 16 patients. Hemodynamic effects of distal ventricular pacing were assessed from the RV apex in all patients, and additionally in a mean of 8 LV sites (range 4-16). LV pacing was achieved via the middle cardiac vein in 10, and another cardiac vein in four. All LVMCO gradients fell with pacing.

The mean LVMCO gradient at baseline was 80±29 mmHg (range 40-139), falling to 31±21 mmHg (range 0-80) when paced from the optimal ventricular site. This represents a mean fall in LVMCO gradient of -49 mmHg (95% CI -62 to -36 mmHg, p<0.0001) (Figure 2). Alternatively, this can be expressed as a mean 60% reduction (range 14-100%). The greatest reductions in LVMCO were from a pacing site in the LV in 14 (87.5%) patients, and from the RV apex in 2.

**Figure 2:**
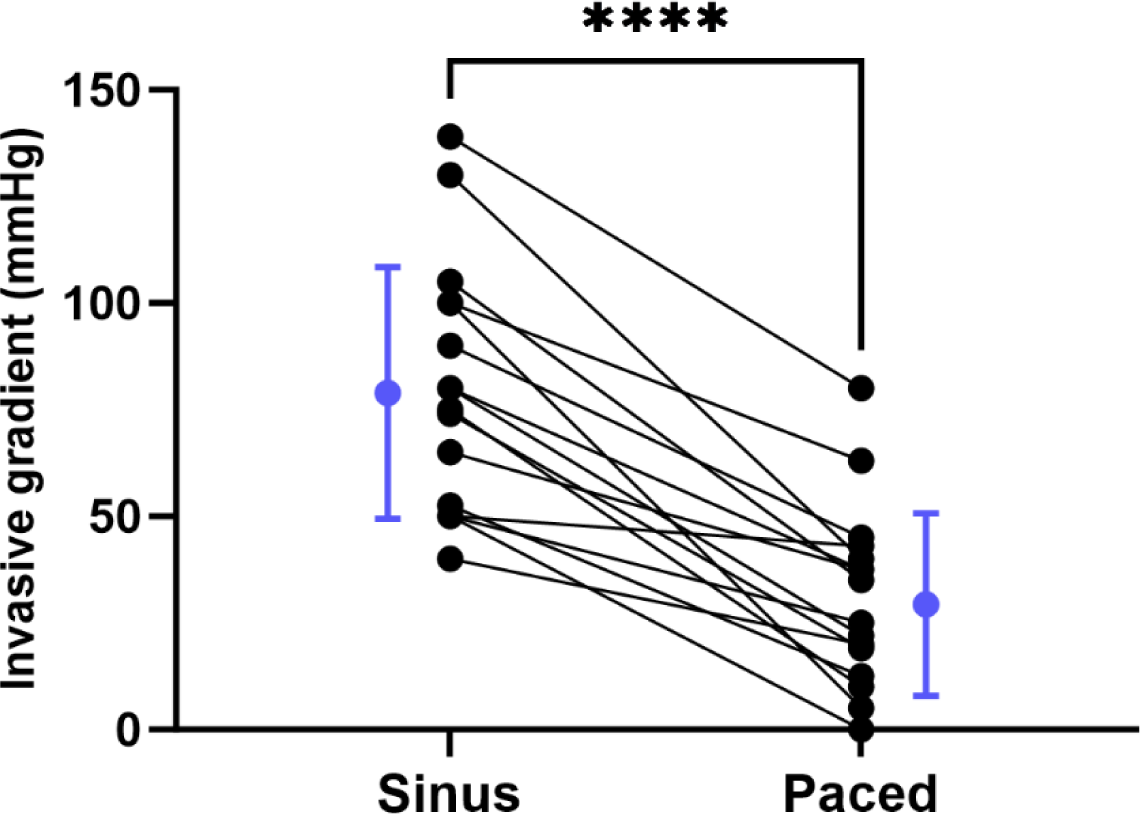
Acute change in LVMCO gradient. Sinus, sinus rhythm; Paced, optimal pacing setting. Error bars: mean±SD. ****=p<0.0001 in pairwise comparisons.

### Secondary endpoints

Pre-implant assessment and both pacing settings during follow-up were completed by 13 patients (with the order of active or back-up pacing randomized) who were eligible for comparison of secondary endpoints between the three time points. Reasons for drop-out are discussed under adverse events below.

### Participant choice of favored pacing setting

At the end of the final study visit, whilst still masked to treatment allocation, subjects were asked in which phase, if any, they felt better. Nine of 13 (69%) subjects chose the active pacing phase as their preferred setting. The remaining 4 (31%) patients reported no difference in symptoms between the two phases.

#### NYHA class

Subjects self-reported NYHA class on ‘good’ and ‘bad’ days (Figure 3 panels A and B). Median NYHA class on a good day was 3 pre implant, 2 in the active pacing arm, and 3 in the back-up pacing arm (IQRs all 2 to 3, p=0.26). The median difference in NYHA class on a good day between active and back-up pacing was zero (p>0.99). Median NYHA class on a bad day was 3 (IQRs 3 to 4) in pre implant, 3 (IQRs 3 to 3) during active and 3 (IQRs 3 to 4) during back-up pacing (p=0.013). The median difference in NYHA class on a bad day between active and back-up pacing was zero (p=0.42). Subjects were also asked to report the ratio of good to bad days throughout the week as: more good days than bad, more bad days than good, or equal good and bad days. More good than bad days were reported with active pacing, and more bad than good days were reported during backup pacing (supplementary Figure S1).

**Figure 3:**
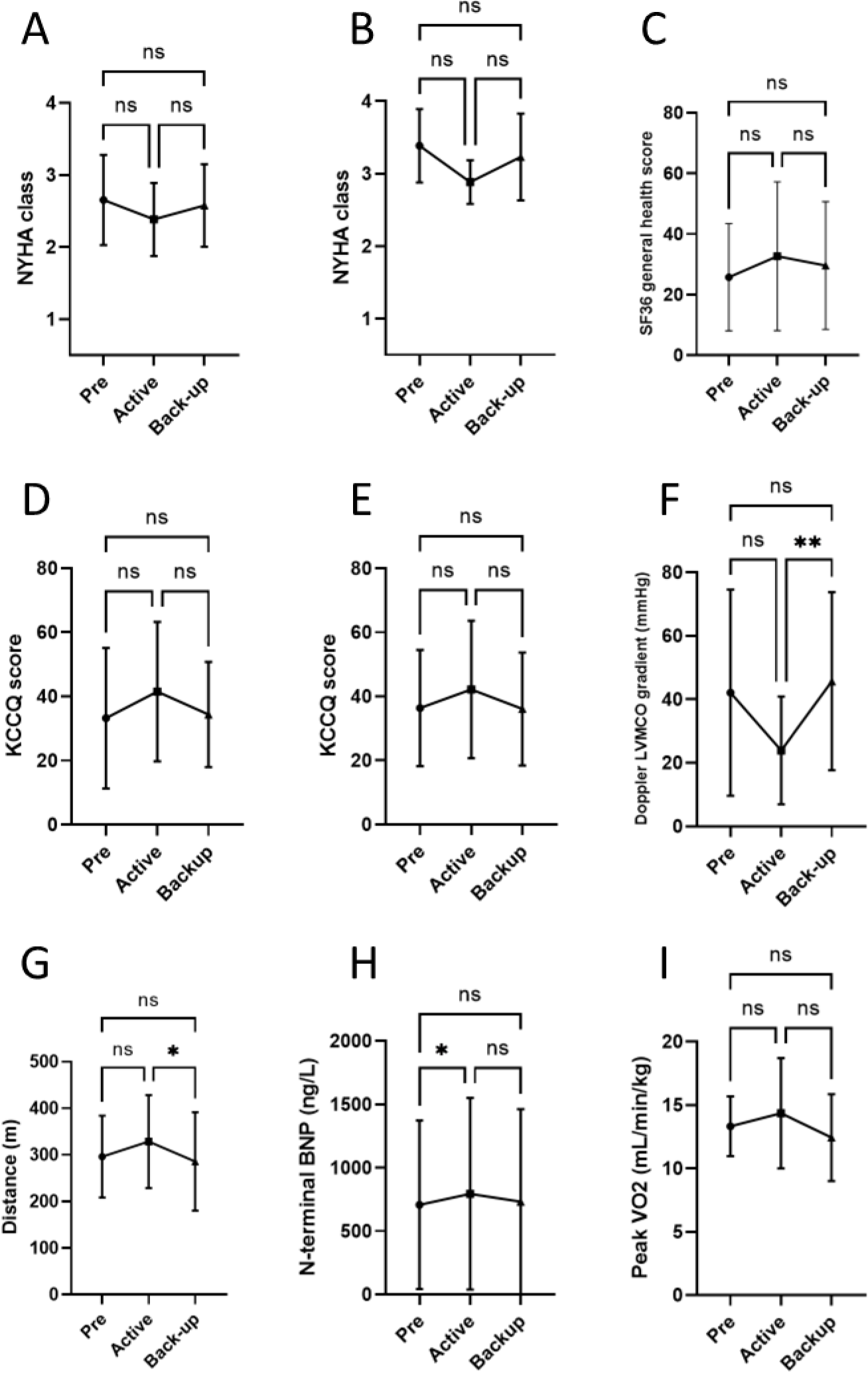
Secondary outcome data across 3 time points: pre-implant, active pacing, and back-up pacing (A-I) (order of active and back-up pacing was randomized). NYHA class on a good and bad day (A & B); SF36 General Health Score (C), KCCQ overall and clinical scores (D & E); Doppler LVMCO gradient (F); 6MWT distance (G); NT-BNP level (H); and VO2 max (I). Error bars: mean±SD. Results of multiple comparisons: ns= p>0.05, *=p<0.05, **=p<0.01 in multiple comparisons.

#### SF36 score

Detailed SF36 results are in Supplementary Figure S2. The mean General Health Score was 26±18 pre-implant, 33±30 during active pacing, and 25±21 during back-up pacing (p=0.18). The mean increase in General Health Score with active compared to back-up pacing was 3 (95% CIs -3 to 9, p=0.42) (Figure 3 panel C).

#### KCCQ score

The mean overall KCCQ score was 33±22 pre-implant, 42±22 during active pacing, and 34±16 during back-up pacing (p=0.22). The mean clinical KCCQ score was 36±18 pre-implant, 42±22 during active pacing, and 36±18 during back-up pacing (p=0.34). The mean increase in overall KCCQ score with active compared to back-up pacing was 7±20 (95% CIs 19 to -5, p=0.44) (Figure 3 panel D). The mean increase in clinical KCCQ score with active compared to back-up pacing was 6±19 (95% CIs 18 to -5, p=0.50) (Figure 3 panel E).

#### Doppler defined LVMCO gradient

The mean Doppler defined gradient was 42±32 mmHg pre-implant, 24±17 mmHg during active pacing, and 46±28 mmHg during back-up pacing (p=0.004). The mean reduction in gradient with active compared to back-up pacing was 21±14 mmHg (95% CIs 11.5 to 29.9, p=0.002) (Figure 3 panel F).

#### 6MWT

The mean 6MWT distance was 296±88 meters pre-implant, 329±100 meters during active pacing, and 286±106 meters during back-up pacing (p=0.038). The mean increase in 6MWT distance with active compared to back-up pacing was 43±47 meters (95% CIs 71 to 14, p=0.018) (Figure 3 panel G).

#### NT-proBNP

The median NT-proBNP concentration was 483 (IQR 243 to 928) ng/L/L pre-implant, 549 (IQR 286 to 1014) ng/L during active pacing, and 422 (IQR 301 to 818) ng/L during back-up pacing (p=0.039). The median increase in NT-proBNP concentration during active compared to back-up pacing was 63 ng/L (IQR 136 ng/L, p=0.12) (Figure 3 panel H).

#### VO2 max

The mean VO_2_ max was 13.3±2.4 mL/min/kg pre-implant, 14.4±4.4 mL/min/kg during active pacing, and 12.4±3.4 mL/min/kg during back-up pacing (p=0.24). The mean increase in VO_2_ max during active compared to back-up pacing was 1.9±4.4 mL/min/kg (95% CIs 1.0 to 4.9, p=0.34) (Figure 3 panel I).

### Safety

#### Hemodynamic pacing study and device implant

16 subjects underwent successful device implantation, of whom 15 (94%) had uncomplicated procedures. Cardiac vein perforation and subsequent pericardial effusion treated with pericardiocentesis occurred in 1 case. In this subject the LV lead was not implanted, and they received a dual chamber ICD without further complication. This subject was withdrawn from the study after the implant procedure.

#### Adverse events during follow-up

There were two patient withdrawals during follow-up due to adverse events unrelated to the study. One patient suffered an acute myocardial infarction, and another went into persistent fast atrial fibrillation in the first follow-up period.

## 4. Discussion

Patients with drug-refractory symptomatic LVMCO have very few therapeutic options.^9, 16^ For the first time, we demonstrate that ventricular pacing tailored to individual patient characteristics is safe, technically feasible, and improves LV intracavity hemodynamics. Furthermore, we present data indicating improved patient functional status, and provide the context and justification for a larger multicenter trial powered for symptom and functional measures.

Other therapeutic options for symptomatic LVMCO include surgical myectomy,^17^ alcohol septal ablation (ASA),^18^ and cardiac transplant.^19^ The novel pharmacological class of myosin inhibitors are not currently licensed for this indication. Prevalence of LVMCO has been reported to be as high as 9-13% of HCM patients.^20, 21^ Notably, as many patients with LVMCO have primary or secondary indications for transvenous ICDs, a trial of PPoC therapy may have a key role earlier in the management of these patients, with progression to other therapies if this fails.

### Surgical Myectomy

Much of the published data on surgery for LVMCO comes from a single center of surgical excellence.^17, 22, 23^ Results indicate similar levels of absolute gradient reduction compared to PPoP. Symptoms were significantly improved with myectomy in a cohort of patients with a less severe phenotype than included in our trial (lower baseline functional limitation and prevalence of LVAA).^17^ Notably, high levels of early complications were reported,^23^ and the surgical expertise required for such specialist procedures is not widely available. By contrast, most centers that implant devices for heart failure already have the experience, expertise and resources required for PPoP. Additionally, as at least half of the published surgical cohorts had pacemakers / ICDs prior to surgery,^17^ a trial of PPoP can be considered before surgery. Finally, unlike for PPoP, no randomized prospective trials of myectomy have been completed.

### Alcohol septal ablation (ASA)

Data on the use of ASA in the treatment of LVMCO is even more limited. ASA reduced obstructive gradients and improved symptoms in a cohort 22 patients.^18^ However, severely elevated residual LVMCO gradients were twice as common when compared to our PPoP cohort (23% vs 12.5%). Furthermore, ASA was associated with intra-procedural complete heart block in a third, and one patient developed ventricular fibrillation.^18^ Once again, there have been no prospective trials of ASA for this indication, and a trial of PPoP prior to ASA may be warranted in the context of baseline ICD indications and the high risk of ASA-related conduction disease.^24^

### Cardiac Myosin Inhibitors

Recent trials of myosin inhibitors, a novel class of agents, report reductions in LV outflow tract obstruction (LVOTO) gradients and improved functional outcomes in patients with ‘classic’ obstructive HCM.^25^ However, perhaps as many as two thirds of HCM patients in EXPLORER-HCM did not achieve primary outcomes, and patients with LVMCO were excluded from the trial. A trial of myosin-inhibitors in symptomatic LVMCO patients is warranted; PPoP may continue to have a role in the management of patients refractory to that treatment, and in those that have indications for transvenous ICDs.

#### Future work

Our trial provides the basis for a larger multicenter trial of PPoP for refractory LVMCO by providing positive signals of safety and clinical benefit, and data required for a study powered to detect clinically meaningful improvements in symptoms and functional status.

While technology developed for resynchronization pacing has enabled this study, there is a need for equipment and techniques developed specifically for this indication. Broadly, these will address challenges that include: predicting optimal lead positioning prior to the invasive procedure; selective intubation of, and attainment of lead stability in the cardiac vein of choice; mitigating effects of myocardial fibrosis on pacing thresholds; and avoiding phrenic nerve capture. Most notably, even small differences in pacing site location can result in strikingly different hemodynamic effects (Figure 4); pre-implant techniques that predict where pacing is likely to have greatest beneficial hemodynamic effects, and an ability to pace beyond anatomical restrictions imposed by cardiac venous anatomy will be key developments.

**Figure 4:**
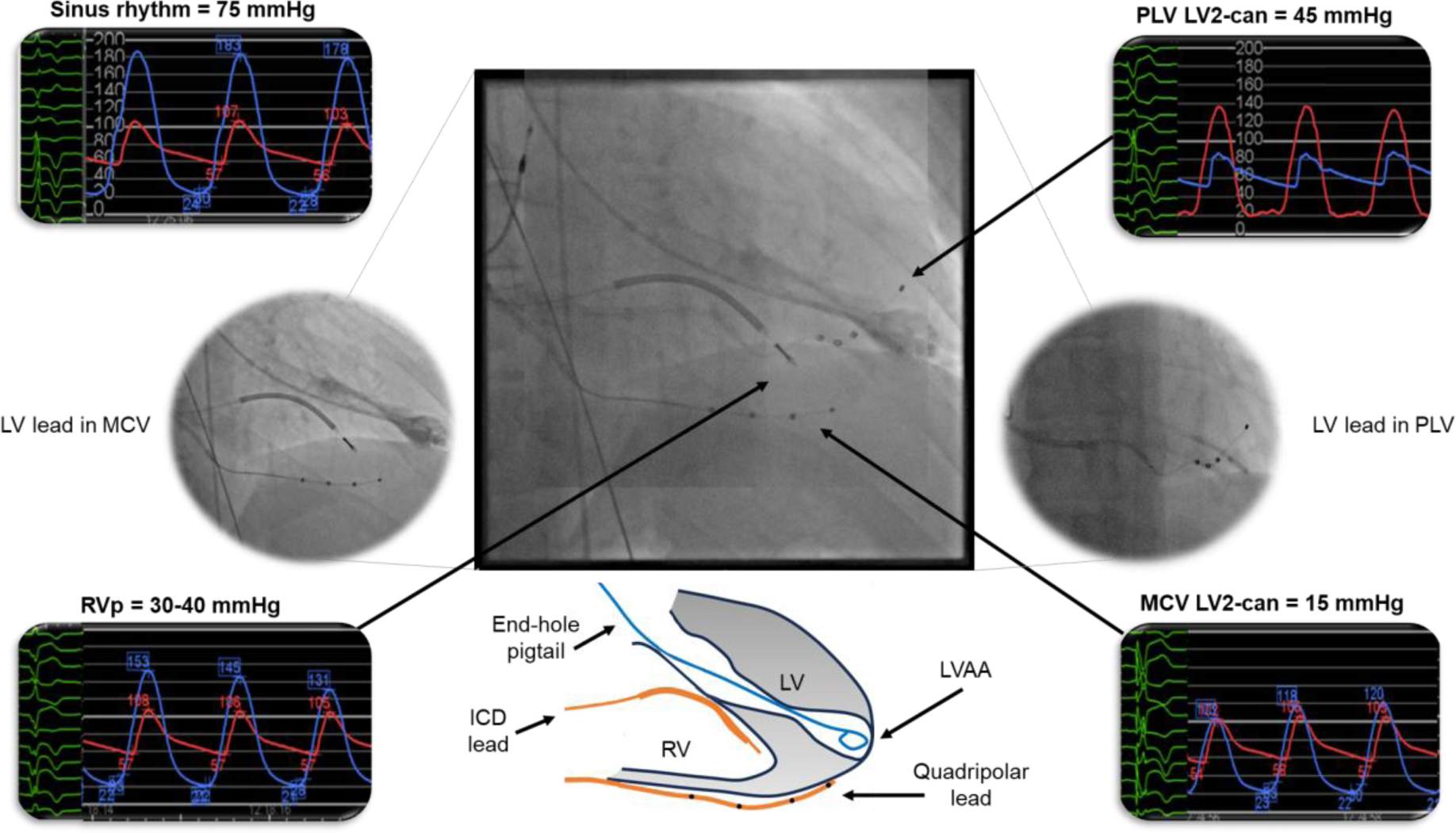
Example pressure traces from hemodynamic pacing study, with overlaid fluoroscopy of the LV quadripolar lead in two different cardiac veins. Pacing from the more lateral position in the PLV produced an unsatisfactory reduction in LVMCO gradient, whereas pacing from the MCV almost entirely abolished the obstructive gradient. A schematic representation of pacing leads and catheter orientation relative to ventricular chambers can be seen at the bottom of the figure. ICD, implantable cardioverter defibrillator; LV, left ventricular; LVAA, left ventricular apical aneurysm; LV2, quadripolar lead pole 2; MCV, middle cardiac vein; PLV, posterolateral vein; RA, right atrial; RVp, right ventricular pacing.

#### Limitations

This study was not powered to detect functional or symptomatic benefit and included only the most severely symptomatic patients. Despite this, we demonstrate significant improvements in the 6-minute walking test, and the overwhelming majority of other functional parameters indicate trends for symptom benefit. Further, to avoid exposing participants to the risks of repeat invasive procedure during follow-up, we relied on Doppler echocardiography to report relative changes in LVMCO gradient, with known shortcomings in this population.^4^ Nonetheless, the significantly lower Doppler-derived gradients during active pacing indicate that the beneficial hemodynamic effect is sustained. Although not significant, mean NT-proBNP was greater following pacing, and was the only secondary outcome not to show a trend towards benefit. Altered atrio-ventricular coupling and/or contractile desynchrony may affect the production of NT-proBNP independently to the magnitude of mid-cavity obstruction; further investigation is warranted.

## 5. Conclusions

In the first randomized placebo-controlled trial of therapy for symptomatic mid-cavity obstructive HCM, we demonstrate that PPoP is a safe and effective therapeutic option. Personalized approaches to pacing most commonly identifies pacing from a site in the LV as the most effective place from which to obtain gradient reduction. Future work will include trials designed to detect symptom and physical performance benefit and attempt to determine how pacing contributes to LVMCO management algorithms that include myosin inhibitors and other invasive therapeutic options.

## Data Availability

Data is available on reasonable request to corresponding author

## Non-standard Abbreviations and Acronyms

AE: Adverse Event
ASA: Alcohol septal ablation
AF: Atrial fibrillation
AV: Atrioventricular
CPET: Cardiopulmonary exercise test
CIs: Confidence intervals
CMR: Cardiac magnetic resonance
CVCTU: Cardiovascular Clinical Trials Unit
ECG: Electrocardiogram
eGFR: Estimated glomerular filtration rate
ESC: European Society of Cardiology
GA: General anesthesia
HCM: Hypertrophic cardiomyopathy
HTN: Hypertension
ICD: Implantable cardioverter defibrillator
IQR: Interquartile range
ISRCTN: International Standard Randomized Controlled Trials Number
KCCQ: Kansas City Cardiomyopathy Questionnaire
LA: Left atrial
LGE: Late gadolinium enhancement
LV: Left ventricle
LVAA: Left ventricular apical aneurysm
LVEF: Left ventricular ejection fraction
LVMCO: Left ventricular mid cavity obstruction
LVOTO: Left ventricular outflow tract obstruction
LVWT: Left ventricular wall thickness
MCV: Middle cardiac vein
MI: Myocardial infarction
NIHR: National Institute of Health and Care Research
NSVT: Non-sustained ventricular tachycardia
NT-proBNP: N-terminal pro B-type natriuretic peptide
NYHA: New York Heart Association
PPM: Permanent pacemaker
PPoP: Personalized prescription of pacing
RV: Right ventricular
SCD: Sudden cardiac death risk
SD: Standard deviation
SF-36: Short form 36 questionnaire
6MWT: 6-minute walk test

## Acknowledgments

The authors would like to acknowledge the contributions of the Data Safety Monitoring and Trial Steering Committees, especially the patient representatives, for their expertise and time.

This study was supported by the Barts Cardiovascular Clinical Trials Unit (CVCTU), a branch of the Barts CTU UKCRC Reg No. 4. The authors would particularly like to acknowledge the work of Anna Bellin, Jackie Cooper, Jane Field, Thomas Godec, and Dilveer Singh Sually for their time and support in the design and running of this trial.

